# Developing a Motherhood regret scale: An examination of reliability, validity, and association with basic attributes

**DOI:** 10.1101/2024.03.13.24304242

**Authors:** Ranno Haruyama, Koubun Wakashima, Kohei Koiwa

## Abstract

The first objective of this study was to develop a scale that measures the level of regret of becoming a mother among women at various stages of motherhood (child age ranging from unborn to 29 years old) and examine its reliability and validity. The second objective was to explore the effects of participants’ basic attributes on the regret of becoming a mother. The study recruited mothers who were pregnant (before birth), mothers of infants (age 0–1), young children (age 2–6), primary school-aged children (age 7–12), and adolescents and young adults (age 13–29). In all, responses from 462 participants (458 women, 4 others; mean age = 37.77 [*SD* = 7.03]; age range = 20–59) were analyzed. As a result, the unidimensional nine-item scale showed an acceptable value for goodness of fit and high reliability (α = .96). In addition, we determined the cut-off score to categorize the regret group. Among the participants in the present study (*N* = 462), 31 were classified as the regret group, and they represented the scores at the top 7%. Moreover, regarding the association between the participants’ basic attributes and level of regret, we found that “having many children” had a significant negative effect on the predicted level of regret (β = –.19, *p* < .001) while “being a single mother” had a significant positive effect (β = .11, *p* < .05). Based on these results, we discuss the structure of the scale that was developed and the effect of a mother’s attributes on the level of regret to provide insights on the attributes of respondents classified into the regret group.

## Introduction

The declining birthrate in Japanese society is becoming increasingly worse. The factors contributing to the declining birthrate are many, including “changes in values toward marriage and giving birth,” “increase in the feeling of responsibility and burden of parenting,” “late marriage, increase in the average age of childbirth, and increase in the number of unmarried people,” and “financial anxiety related to securing funds to cover child-rearing costs” [1,2]. In addition, psychological problems surrounding mothers are on the rise, such as mothers in the child-rearing period becoming more vulnerable to isolation due to the increase in nuclear families brought by the rapid decline in birthrates, the increase in undesired births, postpartum depression associated with parental anxiety, and maternity neurosis caused by the responsibility and pressure of child-rearing [3–6]. Against such trends, the feeling of “regret of becoming a mother” has received more attention in recent years [7].

### The regret of becoming a mother

The “regret of becoming a mother” is defined as the inner feeling of regretting becoming a mother [7,8]. In general, the term “regret” is used to indicate the feeling of disappointment over a choice that was not taken at a point in time between the past and present. In contrast, the “regret of becoming a mother” is different on the point that it is an emotional response expressed between the past and present, and the real and imagined [7]. Thus, in this study, we define the “regret of becoming a mother” as the repentant emotional response that emerges between the reality of being a mother and the imaged reality of not being a mother. In our interpretation, this emotional response is not only recognized against the past, when the decision to become a mother was made, but potentially also in the present of being a mother. Moreover, the term “becoming a mother” as we use it in this study refers to the reality of having a child rather than the process of psychological preparation related to whether the mother herself has or does not have awareness of being a mother.

Donath [7] reports that today, many women around the world live with this regret. In Japan, NHK [9] conducted a survey with a wide range of mothers in their teens to their 70s; one in three people responded that “there has been a time when they wished they had never become a mother” and that they were aware of their regrets and were worried, but they “cannot talk about it (55%),” reflecting a social environment in which mothers cannot attain help. This suggests that many mothers struggle with emotions that they cannot be open about in the current society and culture but are striving to adapt to the role as mothers. Moreover, the hidden increase in the number of mothers in such a situation contributes to the lack of opportunities to share ways to cope with feelings of regret and to the decline in the appeal of becoming a mother, thus maintaining a vicious cycle that the leads to a low-birthrate society.

Therefore, it is necessary to focus on the feeling of “regret of becoming a mother” that is held by many mothers and devise ways to support them. However, current studies on motherhood regret (hereafter, the field) have been limited to sociological accounts of the existence and current situation of regretting mothers [8], and no study has addressed motherhood regret from a clinical psychological perspective for psychological support. Moving forward, it is essential that a support model be presented on how a regretting mother can psychologically and socially adapt to the role of being a mother while having such feelings.

### Factors related to “regret of becoming a mother”

To propose ways to support those experiencing regret of becoming a mother, it is necessary to understand what factors are related to regret. Factors related to motherhood regret that can be inferred from previous studies include parenting anxiety and stress. Matsumura et al. [10] report that the lack of time that a mother can spend on herself influences parenting stress, leading to an increase in the burden of child-rearing. Additionally, Noguchi et al. [11] and Shimizu et al. [12] report that children’s unreasonable behavior accompanied with the sense of not being able to control are contributing parenting stressors. Conversely, studies reporting on the positive side of the topic at hand have shown that having multiple people to talk to can reduce parenting anxiety among mothers of young children and that experiencing growth as a parent and the growth of one’s child are factors that increase the sense of happiness in being a parent [13,14]. From the above, it is predicted that the attributes of a mother influence the factors associated with motherhood regret. Thus, it is necessary to examine such relationships broadly.

### Trends and issues in previous studies

As mentioned above, trends in previous studies dealing with motherhood regret can be understood as those following a sociological perspective and those following a psychological perspective. In the sociological school of thought, the interest has been on pointing out the social pressure that inhibits people from expressing the regret of becoming a mother and pursuing the creation of a society that is tolerant of the diversity of choices related to childbirth and of mothers who reveal their regrets [7,8]. However, in Japan, only a few psychological studies have approached the issue at hand from a psychological perspective, and it is not clear what factors are associated with the mothers’ regret. As a next step, it is important to clarify the factors related to motherhood regret and examine how it is possible for a regretting mother to adapt socially and psychologically as a mother while having a sense of regret.

We identify two problems in the current state of research. First, most of the research on mothers and child-rearing is skewed toward studies targeting mothers of infants. Second, it is not possible to measure the degree of motherhood regret because studies have only measured it by a singular method, such as asking “whether a person regrets [the decision of becoming a mother] or not.” It is expected that a mother’s regret fluctuates along the child’s growth stages and that such fluctuation has continuity. Moreover, it is necessary to measure regret as a matter of degree to examine the factors associated with motherhood regret. Hence, it would be meaningful to develop a scale that can measure the level of regret of becoming a mother by means of question items that target mothers at various child-rearing stages from pregnancy to having adult children.

As such, the two objectives of this study are as follows. The first is to develop a scale that measures the level of regret of becoming a mother among women at various stages of motherhood (child age ranging from unborn to 29 years old) and examine its reliability and validity. The second is to explore the effects of participants’ attributes on the regret of becoming a mother.

## Methods

### Procedures

Data were collected in Japan. The participants were 482 mothers with children of various age groups: before birth, infant (age 0–1), young children (age 2–6), elementary school age children (age 7–12) and adolescents and young adults (age 13–29); of these participants, those with missing answers were excluded from the analysis. In regard to mothers of children aged 15 or older, those who had a score of five or less on the level of memory recollection of the child-rearing period were excluded, and participants with invalid responses to the manipulation check item [15] were excluded as well. Thus, excluding 20 responses in all, responses from 462 participants (458 women and 4 others) were analyzed. The mean age of the participants included in the analysis was 37.77 (*SD* = 7.03, *Range* = 20–59). Participants were recruited using an online service provided by a major Japanese crowd sourcing service administered by Yahoo Japan Corporation (https://crowdsourcing.yahoo.co.jp/). This online service matches crowdsourcing workers to requests from clients. For the present study, we posted a Google Forms questionnaire for the purpose of psychological research in the “survey” category of the online forum. The collection of responses was terminated when the number of respondents reached 130 at each applicable time period or at the end of the posting period. We set the target number of responses so that the sample size would be 50 or more for each applicable time period and exceed the minimum sample size (100 or more responses for analysis) required by the analysis software used in this study: IBM SPSS Amos version 2 and IBM SPSS Statistics version 22 [16,17]. Participants were provided with an electronic informed consent at the time of their participation in the survey and were informed beforehand that they could leave the survey at any time. Consent was obtained with the submission of the participant’s survey responses. Participants who responded to the survey received 55 Japanese yen (approximately 0.5 US dollars) per each submission through the online service. Responses were collected between June 29, 2023 and July 5, 2023.

## Measures

### Sociodemographic variables

Respondents were asked to answer open-ended questions about gender and age, and this was followed by multiple choice questions about the respondent’s attributes. Here, we asked questions that were the same as Donath [8] and that focused on the children’s age group, the number of children, the age of the mother at the time of giving birth, whether they are also a grandmother or not, nationality, religion, highest level of education attained, employment status, marital status, whether they are a single parent, and gender identity. Then, respondents were asked to answer an open-ended question about family composition (their children’s gender and birth order). These items were used to examine the influence of a mother’s attribute on the degree of regret. Hence, to quantify each attribute item, we assigned numerical values to each multiple-choice item in the order in which they were presented. For example, in the question about the “child’s age group,” a higher number indicates that the respondent is a mother to a child in a higher age group. For most or most items, we provided the answer option of “other” and assigned the value “0” to it. However, even if the answer is “Other,” there are cases in which “0” is not assigned. In addition, to quantify the “employment status” item, we based the scale on the degree of involvement in out-of-the-house labor and assigned a score of 1 to “housemaker,” 2 to “seasonal part-time staff,” 3 to “student,” 4 to “contracted labor (fixed-term), ” and “Temporary employment,” 5 to “self-employed,” 6 to “long-term part-time employee,” and 7 to “full-time employee.” Occupations to which the participant did not fall (e.g., internships) were not assigned a numerical value. All participants in the present survey were Japanese nationals. Please refer to the support file for the scoring of these questions and variables (<u>S1 Data</u>).

### Measurement of the degree of regret of becoming a mother

This is a scale item developed for this study. First, we referenced the interview conducted by Donath [8] and the narrative about the feeling of regret experienced by people who regret becoming a mother to select the language for the scale items. Then, we referenced Ueichi’s [18] Japanese translation of Washburn’s regret scale that measures regret proneness in daily life to select and revise the items’ language so that they reflected the feeling of regret as a mother. We revised and edited these with the help of two researchers and faculty who are certified public psychologists and clinical psychologists, obtaining 23 items. This regret of becoming a mother scale is answered on a 5-point Likert-type scale (*strongly disagree* = 1 to *strongly agree* = 5); the higher the score, the stronger the feeling of regret (<u>S1 File</u>).

### Screening of participants who regret becoming mothers

This was the item used in Donath’s [8] interview study to select participants who regretted becoming mothers. In the present study, we used this for the validity test and to define the cut-off value. Below, we explain the screening and statistical procedures. Donath [7,8] used a two-step procedure for screening. As the first step in the present study, this consisted of a maximum of four items; however, it first required a response to the following three questions: (1) do you regret becoming a mother? (2) if you could go back in time and had the knowledge and experience you have today, would you still choose to become a mother (reverse scoring)? (3) in your opinion, does being a mother have any advantages (reverse scoring)? These were all dichotomous questions that were answered with a “Yes” or “No.” As the second step, those who answered “Yes” to item (3) above, that is, those who responded that “being a mother has advantages” were asked to answer an additional item: (4) in your opinion, do the advantages outweigh the disadvantages (reverse scoring)? This was also a dichotomous question answered with a “Yes” or “No.” Based on the responses to these screening items (responses to up to four items), participants were selected if they indicated “regret” in their responses in three or more items. In the present study, we quantified “Yes” answers with a score of 2 and “No” answers with a score of 1 and then processed the reverse scoring items. Next, following Donath’s [8] screening criteria, statistical processing was performed so that a score of seven or above would identify the respondent as eligible for participation. Respondents who had already passed the screening based on their answers to three items out of the four were not directed to the fourth question. In this case, their answers were processed as indicating “regret” and quantified accordingly to be reflected in their score.

### Measurement of regret propensity

We used Ueichi’s [18,19] Japanese translation of Washburn’s regret scale. This scale was designed with the purpose of measuring the extent to which one tends to experience regret [19]. It comprised one factor and 20 items that asked about the degree of regret in daily life situations on a 5-point Likert-type scale (*disagree* = 1 to *agree* = 5); the higher the score, the higher the propensity to experience regret. In this study, we used this for validity check and assumed that the score of this item showed a positive correlation with the degree of regret of becoming a mother.

### Measurement of parenting happiness

We used the short version of the Parenting Happiness Scale designed by Shimizu et al. [20], which measures the degree of happiness gained from parenting among mothers of infant children. This scale comprised 13 items and three factors: five items relating to the “joy in parenting” factor that measures the overall joy a mother experiences in day-to-day parenting, four items relating to the “bond with children” factor that measures the feeling of relief a mother experiences from the realization that the mother–child relationship is unshakable through events that occur between the child and mother themselves or others, and four items relating to the “gratitude towards the husband” factor that measures gratitude for the help with childcare obtained from the husband and others. Items about the happiness a mother experiences in parenting were answered on a 5-point Likert-type scale (*disagree* = 1 to *agree* = 5); a higher score indicated a higher level of happiness in parenting. In the present study, we used the total score of the three factors to check for validity and assumed that the score of this scale showed a negative correlation with the degree of regret of becoming a mother.

### Item to check the accuracy of recalled memory

We used the parenting happiness scale short form designed by Shimizu et al. [20], which was developed specifically for mothers of infant children. Therefore, participants whose children were aged 15 or older were instructed as follows to check the accuracy of recalled memory. Specifically, the instruction message was “The previous questions included items related the care of infants and young children, so please tell us how well you were able to recall those times” and asked the respondent to answer on a 10-point Likert-type scale (*I could not recall at all* = 1 to *I could recall well* = 10). Respondents who had a score of five or less in this check item were deemed as not having provided a valid response to Shimizu et al.’s [20] scale and were excluded from the analysis.

### Measurement of views on motherhood

We used Hanazawa’s [21] Motherhood Concept Questionnaire, which asks about the ideals and values of and attitudes toward motherhood. It comprises 27 items, including items that affirm motherhood (18 items), such as “Child-rearing is a job that suits women well, so it is natural for women to do so” and “If you do not bear and raise a child, there is no worth in being born a woman.” It also contains items that reject motherhood (9 items), such as “It is better to not have children to enjoy married life” and “It is wrong for mothers to make child-rearing their main reason for living.” Respondents were asked to answer their views on motherhood on a 5-point Likert-type scale (*strongly disagree* = 1 to *strongly agree* = 5). For statistical processing, following Hanazawa [21,22], we reverse scored the reject items (9 items) for quantification; thus, a higher score indicated a more positive view toward being a mother. We used this for validity check and assumed that the score of this item has a negative correlation with the degree of regret of becoming a mother.

### Identifying bad respondents

We used the Instructional Manipulation Check (IMC) designed by Masuda et al. [15], which is a questionnaire designed to screen bad respondents who answer online surveys on the Internet without reading the instructions. The instruction read, “In online surveys on the Internet, there is a problem of respondents who provide false answers or without reading the questions. Therefore, we apologize for the inconvenience but allow us to verify if you have read this message. To confirm that you have read and understood this message, please proceed without answering the following question (do not select any option).” The respondent was asked to click “Next” without answering the question.

### Ethical consideration

Consent to the survey and confirmation of consent was done by responding to an online form (Google Form). After explaining the purpose of the study in the first section of the survey, participants were informed that participation was voluntary, that the survey was anonymous, and that personal information would not be disclosed to any third parties. The survey form was designed so that only participants who agreed to these informed consents could participate in the study. thus, consent for participation was considered obtained from respondents once they had completed and submitted their answers. Response records are designed so that individuals cannot be identified for any reason. However, it is possible to print only the record of consent to the research if necessary. This study was approved by the Tohoku University Graduate School of Education’s ethics committee (Approval 23-1-006).

## Data analysis

The analysis was performed using the statistical software IBM SPSS AMOS version 22 and IBM SPSS Statistics version 22. First, we conducted a factor analysis of the motherhood regret scale and a correlation analysis to examine its reliability and validity. Next, we defined the cut-off value to apply to the motherhood regret scale using the index of the completed scale and the index of Donath’s [8] screening items. Finally, we performed a multiple regression analysis to examine the effect of the participants’ attributes on the degree of regret of becoming a mother.

Below, we explain the factor analysis. This scale was assumed to represent one factor. Thus, we conducted a single factor confirmatory factor analysis (CFA) to select the items. Here, we used oblique rotation and maximum likelihood extraction. To verify the goodness-of-fit, we analyzed the goodness-of-fit index (GFI), the adjusted goodness-of-fit index (AGFI), the comparative fitting index (CFI), and the root-mean-square approximation error (RMSEA). The cut-off values for acceptable model fit used in this study are as follows: GFI > .90 as acceptable fit, > .95 as good fit, AGFI > .90 as acceptable fit, > .95 as good fit, CFI > .90 as acceptable fit, > .95 as good fit, RMSEA < .10 as acceptable fit, and < .06 as good fit. In the case of insufficient goodness-of-fit, it was assumed that the error correlation was related to the modification index (MI), which indicates the expected change in parameters when a particular specification is included in the model. In other words, in this study, we interpreted that in a model that measures a single concept, when a model specification that causes a high correlation between items occurs, this is reflected in the value of MI. Therefore, when the degree of fit was insufficient, the analysis was performed based on the MI.

Next, to assess reliability, we calculated the Cronbach’s Alpha Coefficient (α). We followed Oshio’s [23] standards and defined an alpha coefficient of α > .70 as acceptable internal consistency reliability and α > .80 as a high internal consistency reliability.

To assess validity, we calculated Pearson’s correlation coefficient between the motherhood regret scale parameters and the parameters of other scales. We followed the standards set by Cohen [24] to assess validity and interpreted a coefficient of *r* = ±.50 as strong, *r* = ±.30 as medium, and *r* = ±.10 as weak.

Then, we considered the cut-off value to apply in the scale being developed here. We calculated the average score of the motherhood regret scale of screened participants in Donath’s [8] study. We expected a difference between the proportions of screened regretful participants in Donath’s [8] study and those of the participants who met the cut-off value set for the present scale. Thus, instead of using the minimum score in our motherhood regret scale of Donath’s screened participants, we decided to calculate their average score and use this as the feasible cut-off value. Moreover, the NHK study reported that around 30% of mothers had experienced regret [9]; therefore, we considered the proportion of mothers who met the cut-off value for each motherhood stage group.

Finally, to examine the effect of the participant’s attributes on the degree of regret of becoming a mother, we performed multiple regression analysis, in which each attribute item was an independent variable and the degree of regret for becoming a mother was the dependent variable. The stepwise method was adopted as the analysis method because it has the characteristic of specifying only the independent variables that show a significant explanation rate for the dependent variable from among many variables. We assessed multicollinearity following Koshio [23] and confirmed that the variance inflation factor (VIF) was less than 10.

## Results

### Transforming variables into scores and assessing reliability and validity

Table 1 shows the correlation between the basic statistics for each variable measured and the scale.

**Table 1.** Correlation between the Regreting motherhood scale and Other variables.

|  |  |  |  |  |  |
| --- | --- | --- | --- | --- | --- |
|  | Mean (SD) | Min - max | Skewness | Kurtosis | Regreting motherhood scale |
| Regreting motherhood scale ($\alpha=.96$) | 1.62 (.87) | 1.00 - 4.89 | 1.55 | 1.65 | - |
| Donath's screening point ($\alpha=.74$) | 1.12 (.24) | 1.00 - 2.00 | 2.09 | 3.80 | .55** |
| Tendency to regret ($\alpha=.78$) | 3.02 (.50) | 1.40 - 4.60 | -.04 | .20 | .30** |
| Childcare happiness ($\alpha=.92$) | 4.18 (.73) | 1.23 - 5.00 | -1.17 | 1.32 | -.60** |
| Motherhood vision ($\alpha=.90$) | 3.09 (.49) | 1.11 - 4.26 | -.54 | 1.16 | -.53** |
| Notes:**$p<.01$; $\alpha$,Cronbach's alpha. | | | | | |

### Degree of regret of becoming a mother

This is a scale item created for this study. Based on the confirmatory factor analysis and validity test, this scale comprises one factor and nine items. Cronbach’s α was calculated to assess reliability, and an α coefficient of .96 indicates high reliability as it sufficiently meets the criteria of α > .80 of high internal consistency reliability.

### Screening items for those who regret becoming mothers

This corresponds to the four items used in Donath’s [8] study to select mothers who experienced feelings of regret at the time of the interview. These were all “Yes” or “No” dichotomous questions and given a score of Yes = 2 and No = 1. The α for the total score was .74, which met the α > .70 criteria for acceptable internal consistency, and the reliability of the scale was confirmed. We used this to assess the validity of the motherhood regret scale.

### Regret propensity

This is a 20-item scale that measures the propensity of experiencing regret in daily life. The α for the total score was .78, which met the α > .70 criteria for acceptable internal consistency, and the reliability of the scale was confirmed. We used this to assess its validity.

### Parenting happiness

It is a 13-item scale that measures the degree of happiness that a mother experiences through parenting. The α for the total score was .92, which met the α > .80 criteria of sufficient internal consistency, and high reliability was confirmed. We used this to assess the validity of the motherhood regret scale.

### View of motherhood

It is a 27-item scale that measures the ideals and values of and attitudes toward motherhood through positive and negative views of it. The α for the total score was .90, which met the α > .80 criteria of sufficient internal consistency, and high reliability was confirmed. We used this to assess the validity of the motherhood regret scale.

### Validity assessment

We calculated the totals of the regret scale score, Donath’s [8] screening items score, regret propensity score, parenting happiness score, and view of motherhood score and calculated the correlation coefficients for each. The results show a significant correlation at the 1% level between each of the variables (Table 1). Specifically, the results showed a significant positive correlation between the degree of regret of becoming a mother and the screening item score (*r =* .55, *p* < .01) and a positive correlation between the degree of regret of becoming a mother and regret propensity (*r* = .30, *p* < .01). Furthermore, the degree of regret of becoming a mother had a significant negative correlation with parenting happiness (*r* = –.60, *p* < .01) and a negative correlation with view on motherhood (*r* = –.53, *p* < .01); thus, the motherhood regret scale was found to show sufficient validity.

### Factor structure of the motherhood regret scale

We performed a one factor confirmatory factor analysis of the original 23-item motherhood regret scale. Table 2 shows the model fit comparison (original 23 items; Table 2, Model 1). In the process of repeating factor analysis, we excluded the items for which the factor loading did not reach .80. In addition, we hypothesized the relationships between items as follows: a positive correlation between item 7 (“I feel being a mother is unbearable”) and item 8 (“if I could, I would like to quit being a mother”), a negative correlation between item 9 (“I believe that I will never be happy as a mother”) and item 16 (“I sacrificed myself to become a mother and this left a scar on me”), and a negative correlation between item 10 (“I believe a life in which I do not become a mother would had been better”) and item 19 (“Becoming a mother was an event that will continue to hurt me throughout my life”). Thus, by drawing a pass based on the modification index, we were able to attain an acceptable degree of goodness-of-fit, and an acceptable degree of consistency was confirmed (χ^2^ = 111.69, *p* < .001, GFI = .95, AGFI = .91, CFI = .98, RMSEA = .09 [90% Cl = .07–.11]). Table 2 shows the results of the final factory analysis (9 items after selection; Table 2, Model 2), and Fig 1 shows the model with the acceptable degree of fit.

**Fig 1.**
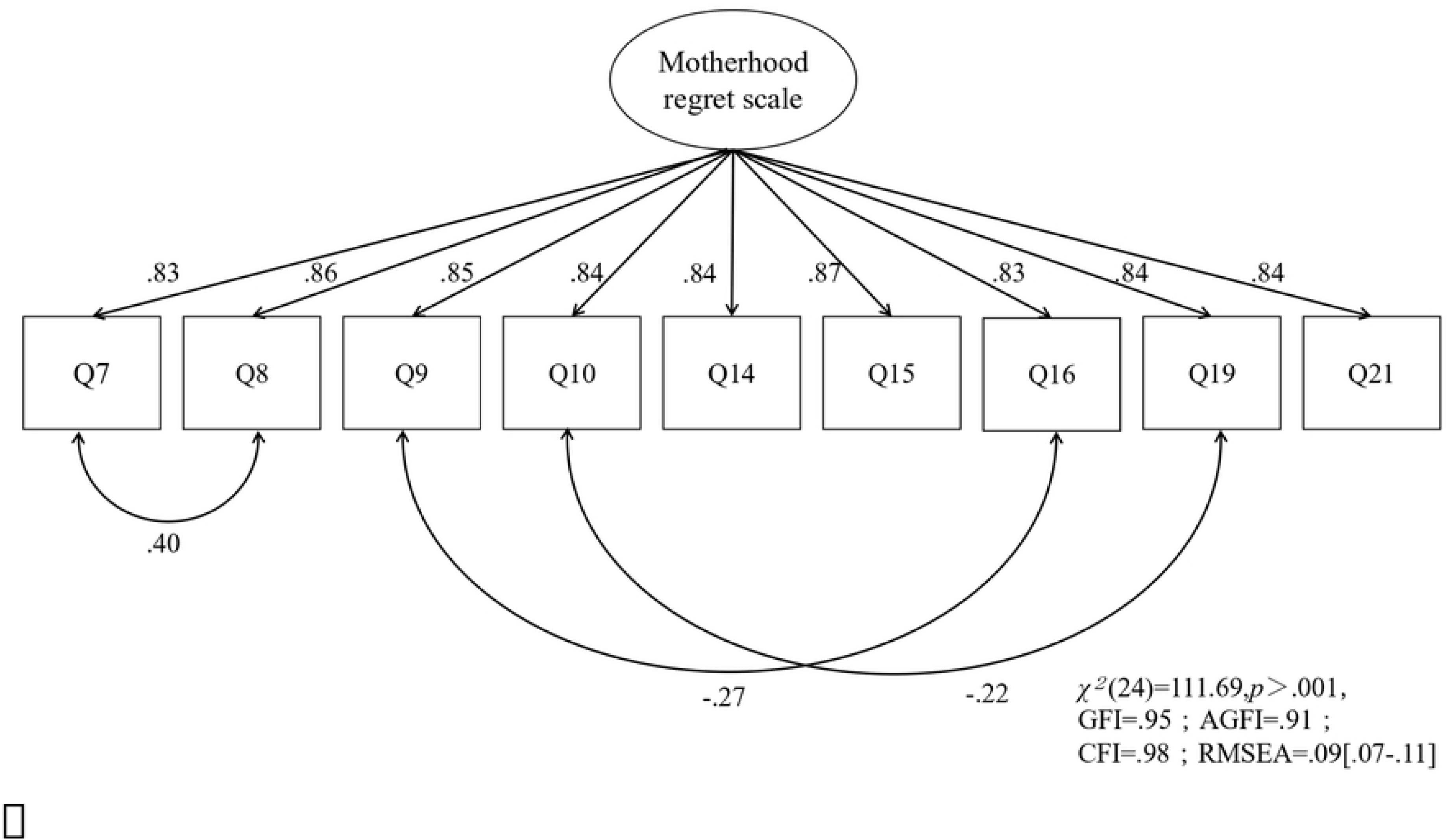
This is the Fig 1 Title. Fig 1. Path diagram of the suitable model.

**Table 2.** Factor loadings for the Motherhood regret scale.

| Table 2. Factor loadings for the Motherhood regret scale. |  |  |  |
| --- | --- | --- | --- |
| Items |  | Standardized loadings |  |
|  |  | Model 1 | Model 2 |
| **Regreting motherhood scale ($\alpha=.96$)** |  |  |  |
| 1 | Think how much better my life would have been if I had chosen not to be a mother. | .72 | – |
| 2 | I think I should have been more careful about whether or not to become a mother. | .72 | – |
| 3 | I think it is unfortunate to be a mother. | .78 | – |
| 4 | I think I became a mother to meet the expectations of society and others. | .63 | – |
| 5 | I believe that it was never my intention to become a mother. | .74 | – |
| 6 | I think I should never have become a mother. | .80 | – |
| 7 | I feel that being a mother is unbearable. | .86 | .83 |
| 8 | I would quit being a mother if I could. | .87 | .86 |
| 9 | I don't think I will ever be happy as a mother. | .81 | .85 |
| 10 | I would have preferred not to be a mother. | .84 | .84 |
| 11 | I feel terrible when I look at my children. | .73 | – |
| 12 | When I look at my children, I don't think they are cute. | .78 | – |
| 13 | I feel traumatized by motherhood. | .79 | – |
| 14 | When I hear the words "women who don't want to have children," I think it is me. | .83 | .84 |
| 15 | When I realize (or reconfirm) that I have become a mother, I feel hazy feeling. | .87 | .87 |
| 16 | I feel as if motherhood has made sacrifices and left scars. | .82 | .83 |
| 17 | When I think of being a mother, I feel like I am in a place where I cannot leave. | .70 | – |
| 18 | I feel trapped by motherhood. | .76 | – |
| 19 | I feel that motherhood was an event that scarred me for life. | .82 | .84 |
| 20 | I feel oppressed by the world's expectations that I should love my children. | .77 | – |
| 21 | Feel that I do not like being a mother. | .85 | .84 |
| 22 | Feel a great deal of frustration at being a mother. | .81 | – |
| 23 | Feel forced to think that being a mother is a joyful thing. | .71 | – |
| $\chi^2$ | | 1926.29*** | 111.69*** |
| df |  | 230 | 24 |
| GFI |  | .66 | .95 |
| AGFI |  | .60 | .91 |
| CFI |  | .84 | .98 |
| RMSEA |  | .13 | .09 |
| 90 Percent C.I. |  | .12-.13 | .07-.11 |
| Notes:***$p < .001$;GFI,goodness-of-fit index;AGFI,adjusted goodness-of-fit index ;CFI,comparative fitting index;RMSEA,root-mean-square approximation error;$\alpha$,Cronbach's alpha. | | | |

### Considerations for the cut-off value and proportion of participants classified into the regret group Cut-off value

We calculated the average motherhood regret score of the screened participants from Donath’s [8] study and used this score as the cut-off value (Table 3). The average score of the screened participants (*N* = 26) was 30.38, and we set the cut-off value for our scale at 30. Therefore, we classified a score of 30 or higher out of a total possible score of 45 into the regret group and a score of 29 or less into the non-regret group. In the current study, 31 of the 462 participants (7%, *Range* = 26–53) had a score that falls under the regret category. In contrast, 13 of the 26 screened participants in Donath’s [8] study were classified into the regret group using our scale.

**Table 3.** Examining the cutoff point.

| Table 3. Examining the cutoff point |  |  |
| --- | --- | --- |
|  | Regreting motherhood scale |  |
|  | Mean [*Range* ] | *N* (%) |
| Donath's screening selector (SI:8点) | 36 [28 - 44] | 12 (3) |
| Donath's screening selector (SI:7点) | 26 [16 - 35] | 14 (3) |
| Donath's screening selector everyone | 30 [16 - 44] | 31 (7) |
| Notes:SI,screening items. |  |  |

### Participants classified into the regret group

The proportion of participants identified as the regret group was 7% (*N* = 31, *Range* = 26–53) of the total sample. The breakdown (Top % of each stage, mother’s age range) by motherhood stage is as follows. Among pregnant mothers, seven out of 64 (11%, *Range* = 27–42), 3 out of 55 mothers of infants (5%, *Range* = 30–36), 11 out of 131 mothers of young children (8%, *Range* = 26–42), 5 out of 133 mothers of elementary school age children ( 4%, *Range* = 35–41), and 5 out of 79 mothers of adolescents and young adults (6%, *Range* = 30–53) were classified into the regret group (Fig 2).

**Fig 2.**
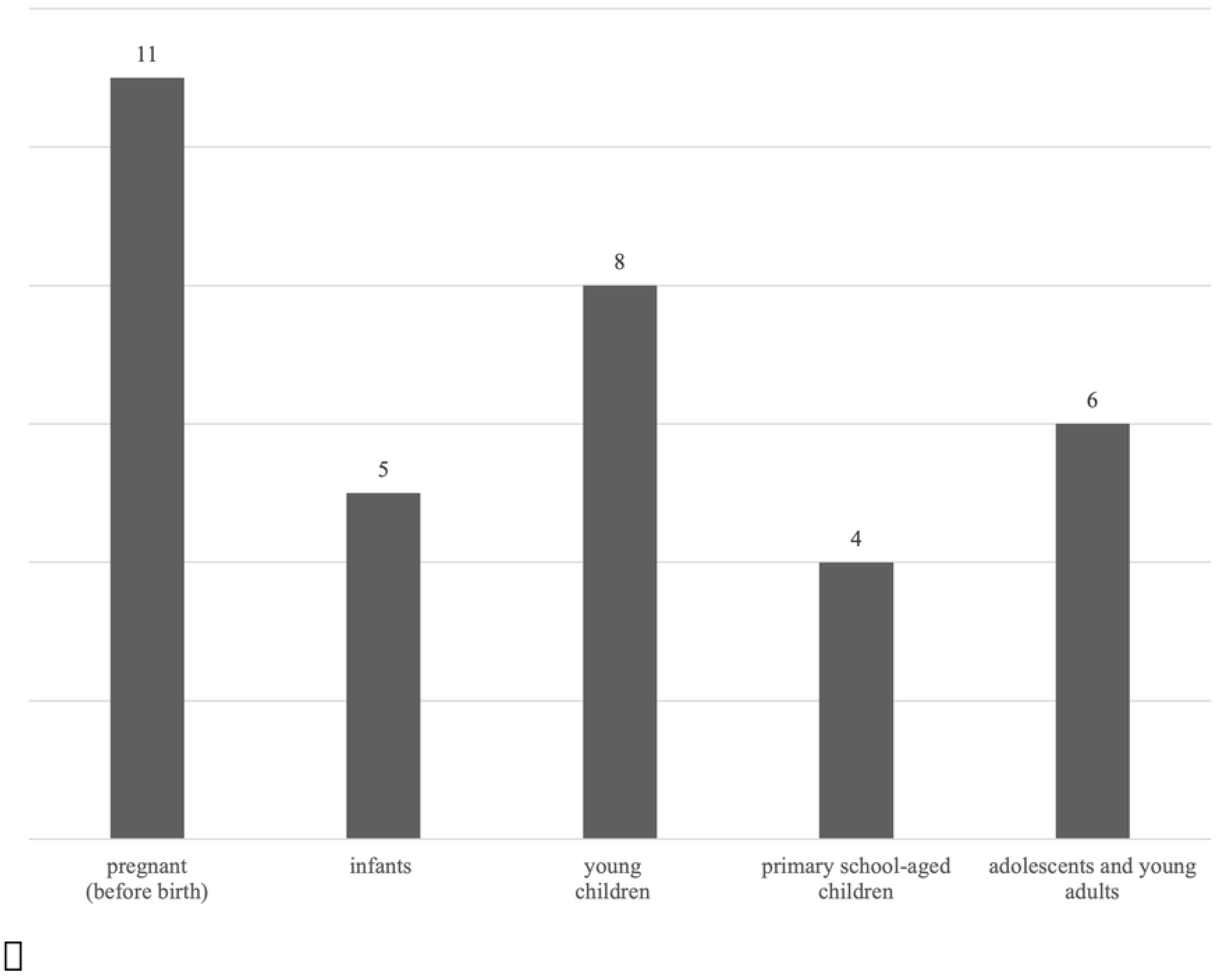
This is the Fig 2 Title. Fig 2. Proportion of mothers classified into the regret group by motherhood stage groups (%)

### The influence of attributes on the regret of becoming a mother

Table 4 shows the results of the multiple regression analysis (stepwise procedure) conducted to understand the participants’ attributes and the influence of such attributes on the degree of regret of becoming a mother. In the analysis, we defined the items for participants’ basic attributes as the independent variables and the degree of regret of becoming a mother as the dependent variable. The variance inflation factor was below 10 for all, and multicollinearity was not observed. The results showed that the attribute “having many children” had a significant negative correlation with the estimated degree of regret of becoming a mother (β = –.19, *p* < .001) and “being a single mother” had a significant positive correlation with the estimated degree of regret of becoming a mother (β = .11, *p* < .05).

**Table 4.** Results of multiple regression analysis using subject attributes and degree of regret over becoming a mother as dependent variables.

| Variable | *N* = 462 | % | *B* | $\beta$ |
| --- | --- | --- | --- | --- |
| Sex |  |  |  |  |
| Male, Others | 4 | (1%) |  |  |
| Female | 458 | (99%) |  |  |
| Age |  |  |  |  |
| ≤20s | 0 | (0%) |  |  |
| 20s | 46 | (10%) |  |  |
| 30s | 229 | (50%) |  |  |
| 40s | 162 | (35%) |  |  |
| 50s | 25 | (5%) |  |  |
| The children's age group |  |  |  |  |
| Pregnant (before birth) | 64 | (14%) |  |  |
| Infant (age 0–1) | 55 | (12%) |  |  |
| Young children (age 2–6) | 131 | (28%) |  |  |
| Elementary school age children (age 7–12) | 133 | (29%) |  |  |
| Adolescents and young adults (age 13–29) | 79 | (17%) |  |  |
| The number of children |  |  | -1.72 | -.19*** |
| 1 person | 231 | (50%) |  |  |
| 2 person | 165 | (36%) |  |  |
| 3 person | 53 | (11%) |  |  |
| 4 or more person | 13 | (3%) |  |  |
| The age of the mother at the time of giving first birth |  |  |  |  |
| ≤20s | 2 | (0%) |  |  |
| 20s | 227 | (49%) |  |  |
| 30s | 222 | (48%) |  |  |
| 40s | 9 | (2%) |  |  |
| 50s | 2 | (0%) |  |  |
| Grandmother | 4 | (1%) |  |  |
| Nationality (Japan) | 460 | (100%) |  |  |
| Religion |  |  |  |  |
| No religion | 417 | (90%) |  |  |
| Belongs to a religion (Buddhism, Shinto, Soka Gakkai, Shinnyoen, Shingon Esoteric Buddhism, Shingon Buddhism, Jodo Shinshu, Catholicism, Christ) | 45 | (10%) |  |  |
| Final education |  |  |  |  |
| Junior high school | 5 | (1%) |  |  |
| High school | 82 | (18%) |  |  |
| Universities, Vocational, Junior colleges | 355 | (77%) |  |  |
| Master's course | 15 | (3%) |  |  |
| Doctor's course | 3 | (1%) |  |  |
| Others | 2 | (0%) |  |  |
| Employment status |  |  |  |  |
| Housemaker (No outside work) | 183 | (40%) |  |  |
| Seasonal part-time staff | 91 | (20%) |  |  |
| Student | 1 | (0%) |  |  |
| Contracted labor (fixed-term), Temporary employment | 19 | (4%) |  |  |
| Self-employed | 23 | (5%) |  |  |
| Long-term part-time employee | 16 | (3%) |  |  |
| Full-time employee | 122 | (26%) |  |  |
| Others | 7 | (2%) |  |  |
| Internship (Paid & unpaid) | 0 | (0%) |  |  |
| Marital status |  |  |  |  |
| Married | 437 | (95%) |  |  |
| Unmarried, Others | 25 | (5%) |  |  |
| Single mother | 22 | (5%) | 3.69 | .11* |
| Gender identity (Female) | 448 | (97%) |  |  |
|  |  |  | *adj R*<sup>2</sup> | .039 |

\**p* < .05, \*\**p* < .01, \*\*\**p* < .001

## Discussion

### Designing a scale

The primary objective of this study was to develop a scale that measures the degree of regret of becoming a mother among women at various stages of motherhood (child age ranging from unborn to 29 years old) and examine its reliability and validity. Below, we discuss the factor structure, reliability and validity of the scale, and cut-off value applied, as well as the characteristics of the item components. For the instrument’s structure, we expected the scale to be unidimensional and thus used a one factor confirmatory factor analysis. After the analysis, the original 23 items were refined into 9, and the model showed acceptable fit indices and high reliability. As for validity, we confirmed sufficient validity by verifying that the regret of becoming a mother scale had significant correlations with each scale as hypothesized. Specifically, the results showed that a high degree of regret of becoming a mother is associated with high propensity to experience regret in general in daily life, low happiness from parenting, and low levels of positive views of motherhood. Therefore, it is presumed that those who regret becoming a mother tend to be more likely to experience regret in other aspects of daily life other than motherhood and cannot have positive feelings about parenting and their role as mothers.

Next, we explain the cut-off point of the scores. A widely used method to determine the cut-off point is the receiver operating characteristic (ROC) curve analysis, which draws an ROC curve based on the reference value of the scale for which the cut-off point is being determined (the scale being tested) and for other equivalent scales (the test scale). Then, for observations that exceed the reference value in the test scale, the minimum value in the scale being tested is calculated, and this is used as the discrimination threshold. On this point, it was expected that the variation in the scores of Donath’s [8] screening items would be large since it is not a statistically tested accurate scale. Hence, it was hard to assume that the scores would be appropriate to set a cut-off value for our scale; in fact, the range of scores for the present scale among Donath’s [8] screened participants were wide ranging, from a minimum score of 16 out of 45 and a maximum of 44 out of 45. Therefore, in the present study, we calculated the average score of Donath’s [8] screened participants and used this to set a cut-off value of 30. Participants who scored above this threshold represented 7% of the total sample, which concurs with the proportion of people that reported regret (7%) in NHK’s [9] report. Thus, it was concluded that we had reached a reasonable cut-off value. In summary, we developed a scale that is unidimensional, with nine items answered on a 5-point Likert-type scale, and with a cut-off value score (30 out of 45) to categorize the regret group.

In terms of item content, we were able to design an item asking about the “regret of becoming a mother” that does not depend on the children’s age-specific events; therefore, we addressed the issue of skewed participant characteristics in previous studies. The commonality among the items created allows to group them into “pressure of being a mother” (items 7 and 8), “despair and hopelessness in being a mother” (items 9, 16, 12), and “negative emotions when objectively recognizing the fact of being a mother” (items 10, 14, 15, 19). We suspect that such feelings are related to the desire to escape from compliance with the socially expected role as a mother and a negative sense toward motherhood itself. On the other hand, reflecting on the process that common item groups such as “negative feeling towards the decision of becoming a mother” (item 1, 2, 4, 5, 6, 13) and “not being able to love my child” (item 11, 12, 20) resulted in being excluded, we suggest that the respondents held a negative view of themselves as mothers, but their regret of the decision in the past is resolved, and the regret they hold is not associated with lack of love toward the child.

### The influence of basic attributes on the degree of regret

A second objective of the study was to investigate the effects of participants’ basic attributes on the regret of becoming a mother. Results of multiple regression analysis with each attribute item as the independent variable and the degree of regret of becoming a mother as the dependent variable showed that “having a large number of children” was a significant negative predictor of the mother’s degree of regret, and “being a single mother” was a significant positive predictor of the mother’s degree of regret. Therefore, the greater the number of children, the lower the mother’s degree of regret, and being a single mother is associated with a higher degree of motherhood regret.

Based on the above result that a greater number of children is associated with a lower degree of regret, it can be argued that when a mother experiences less regret, she is more likely to decide to have another child. From this perspective, it is suggested that another factor leads to the absence of regret against the reality of being a mother. On the other hand, studies in Japan with child-rearing mothers report that as the number of children increases a mother’s concern for not spending enough time with her children and being left with less time for herself [25]. At first glance, such reports suggest that a mother’s regret will be higher due to the large number of children; however, the insights from the present study suggest that children bring some sort of positive effect, and overall, an association exists a greater number of children and a low degree of mother’s regret. Here, considering studies that focus on the mothers’ positive emotions, it is documented that one of the factors increasing parents’ sense of happiness in parenting is when they feel their own growth as a parent or their children’s growth [13]. Thus, it can be inferred that as the number of children increases, the opportunities to experience growth as a parent and joy from the process of parenting also increase, in turn leading to a lower degree of regret. In the future, it will be meaningful to investigate what child-related factors are associated with a low degree of mother’s regret.

The results showing the association of being a single mother with a high degree of regret may be an indicator of the harsh reality of a life as a single mother [26]. Nakasono [27] points out that 80% of single mothers are employed, but at the same time, 50% live in poverty, and they are in a condition of being “working poor.” Relatedly, Matsumura et al. [10] suggest that the fact that mothers have less time to spend on themselves affects parenting stress and leads to an increase in the burden of child-rearing. In this way, the present situation in which mothers spend most of their time in the labor force, caring for the household, and attending to children has not changed much even after the pandemic and its social effects [26–28]. Moreover, spouse participation in parenting increases mothers’ sense of happiness and decreases the burden from child-rearing [2,29]. Furthermore, mothers who build close relationships through interpersonal interactions in the local community experience parenting as fun and thus a lower sense of burden, in addition to showing satisfaction in their daily lives [25]. In summary, the reality is that single mothers have most of their time taken by work, parenting, and other responsibilities as mothers; they have little time for themselves and are missing the opportunity to participate in local community social spaces. Such a reality, in addition to not having the support of a spouse, seems to be associated with high degrees of regret.

### Participants classified into the regret group

Below, we consider the participants who were classified into the regret group and their motherhood stages. Among the respondents classified into the regret group in the present scale (*N* = 31), of those who passed Donath’s [8] screening (*N* = 26), the majority (*N* = 13) scored below the cut-off value of 30 and were excluded from the regret group. In addition, the NHK [9] report indicated that one in three (30%) mothers regretted becoming mothers, but our results show that the proportion was 1 in 14 (7%); these differences could be explained by different methods of measurement. NHK’s [9] report asked participants whether regret had been “experienced at least once in the past,” while our study asked about “mother’s regret recognized in the present.” Thus, many have had regrets for becoming mothers in the past, but not many mothers currently regret this. In other words, even if a mother had once felt regret, she acquired the means to adapt to this regret over time and the skills to escape from regret and strive for the role of mother.

In addition, when examining the data by motherhood stage groups, the proportion of mothers belonging to the regret group (*N* = 31) was the highest. The proportion decreased among mothers of infants and increased among mothers of young children; subsequently, the proportion decreased among mothers of elementary school age children and increased among mothers of adolescents and young adults. According to such trend, mothers’ regret increases between children’s transition from infancy to early childhood and between elementary school age to adolescence/adulthood. What this pattern shows is correspondence with the rebellious phases observed along the developmental stages of a child. In the present study, early childhood is the first rebellious period, and adolescence/adulthood the second; these are periods when a mother experiences changes in her sense of control in parenting. Studies examining sense of control among mothers of infants have reported that children’s unreasonable behavior is a factor that leads to parenting stress in mothers [11] and that the sense of inability to control the children is an element that explains parenting stress in mothers [12]. Therefore, it is suggested that changes in regret are related to a mother’s own sense of control; in fact, during pregnancy, the mother cannot control her own body in the same way she used to before the pregnancy due to the changes in her body. When a child transitions from infancy to early childhood, they start moving around at their own will and verbally expressing their will, making it hard to control their behavior.

Next, in the transition from childhood to adolescence/adulthood, the child becomes more independent psychologically, and the relationship with the mother also changes. Therefore, a mother may experience regret during periods when a change in the sense of control in respect to the mother’s body, the child’s behavior, and the psychological realm occurs. However, since the present study is limited to the measurement of regret, we look forward to future studies examining the factors related to changes in regret.

### Limitations of the study

This study has several limitations. First, we should consider the sample. Although data were collected from a wide range of people using the Internet, it is highly likely that the participants in our study had sufficient time to take part in the survey and were in an emotionally stable condition to answer about the regret of being a mother. Future studies should examine a wide range of factors related to living conditions, such as financial situation, weak ties to supportive networks for performing the role as a mother, and being a single mother, and investigate how these factors influence the regret of being a mother.

Next, only using the motherhood regret scale does not provide insights into the causes and factors related to regret. For instance, the reasons and factors related to regret may be different for mothers who show the same score but are in different stages of motherhood. In the future, it will be necessary to examine what factors constitute regret in each motherhood stage.

Last, although we discussed our thoughts about the proportion of the regret group by each motherhood stage, since we used cross-sectional data, it is not possible to examine how individuals progressed through the changes in regret. In the future, it is recommended to collect and examine data longitudinally though such methods as case studies.

## Conclusion

As stated above, this study was able to explain the factor composition, reliability, and validity of the “Motherhood Regret Measurement Scale” despite its limitation. Moreover, it showed the relevance of the mother’s attribute on the degree of regret. Our study is the first in Japan to focus on motherhood regret from the perspective of clinical psychology and quantitatively demonstrate its relevance.

The body of research on family, parenting, and mothers’ mental health is growing. Against this backdrop, our study contributes to a greater understanding, acceptance, and support of the regret that many mothers face by making it possible to quantitatively measure its degree within a sociocultural context where support is often overlooked.

## Supporting information

S1 File. Motherhood regret scale. (<u>PDF</u>)

S1 Data. Anonymized data set. (<u>CSV</u>)

## Data Availability

All relevant data are available within the manuscript.

## Acknowledgements

We would like to thank everyone who cooperated in the study and composition of this paper.

